# Changes Over Time in COVID-19 Vaccination Inequalities in Eight Large U.S. Cities

**DOI:** 10.1101/2021.12.01.21267158

**Authors:** S. Michael Gaddis, Colleen M. Carey, Nicholas V. DiRago

## Abstract

We estimate the associations between community socioeconomic composition and changes in COVID-19 vaccination levels in eight large cities at three time points. Between March and April, low SES communities had significantly lower change in percent vaccinated than high SES communities. Between April and May, this difference was not significant. Thus, the large vaccination gap between communities during restricted vaccine eligibility did not narrow when eligibility opened up. The link between COVID-19 vaccination and community disadvantage may lead to a bifurcated recovery where advantaged communities move on from the pandemic more quickly while disadvantaged communities continue to suffer.

## INTRODUCTION

In early 2021, state and local authorities in the United States vaccinated millions of individuals weekly against COVID-19. By late April, over 80 million people, about one-quarter of the U.S. population, were fully vaccinated, reducing their risk of symptomatic and asymptomatic infection, transmission, hospitalization, and death.^1–3^ In many urban areas, vaccine doses were scarce through April.^4^ Vaccine eligibility progressed in stages, starting with health care workers and proceeding, per state and local policy, to individuals of advanced age, in certain occupations, or with particular comorbidities.^5^ By May, vaccine supply approached demand in more places,^6^ and 44 states and the District of Columbia had expanded vaccine eligibility to everyone age 16 and older.^7^

We may expect that the response and recovery period of the COVID-19 pandemic has differentially affected individuals and communities based on existing socioeconomic (SES) disadvantage. As others have noted, populations facing disadvantage prior to a major public health crisis fare worst both during and after the crisis.^8^ To date, evidence from the COVID-19 pandemic suggests a similar story. First, neighborhoods and communities with higher levels of socioeconomic disadvantage have been hardest hit during the earliest stages of the pandemic. Incidences of infection and mortality have been higher where low-SES individuals and people of color (POC) comprise more of the population.^9–14^ Second, researchers have documented community inequalities in COVID-19 vaccinations by neighborhood disadvantage during early restricted vaccination eligibility periods.^15,16^

It is unclear, however, how much the recovery of disadvantaged neighborhoods has lagged after restricted vaccination eligibility periods. In this research, we examine whether existing gaps in vaccination rates between advantaged and disadvantaged neighborhoods closed as vaccine eligibility expanded. We examine this issue using vaccination data from eight cities over three time points between March 21 and May 3, 2021, capturing the onset of widespread eligibility. Our findings contribute to a rapidly growing body of literature examining inequities both due to the pandemic and as a result of the response and recovery phases.

## DATA AND METHODS

We gathered official counts of the percentage of individuals with at least one dose of a COVID-19 vaccine by ZIP Codes (hereafter: communities) in eight of the ten most populous U.S. cities: New York, Chicago, Houston, Phoenix, Philadelphia, San Antonio, San Diego, and Dallas. We collected these data at three time points: March 21, April 12, and May 3, 2021. To account for community composition, we used American Community Survey (ACS) data on percent employed in “health care and social assistance” and age 65 and older (to control for early eligibility); percent enrolled in Medicaid or other means-tested public health insurance, without health insurance coverage, under the federal poverty line, and without internet access (to measure socioeconomic status [SES]); percent Black, Hispanic, Asian, and White (to control for race/ethnicity). We converted each SES variable into quartiles standardized by city.

We estimated population-weighted linear regressions using Stata/MP version 16.1. We used the margins command to estimate adjusted predictions at the means and adjusted predictions for low and high SES communities. For the latter two, we set all SES variables to either the lowest or highest quartiles and all other variables at their means. Our analyses provide the adjusted regression estimates for percent of vaccinations overall, in low SES, and in high SES communities in March, April, and May, and changes in vaccinations over time.

We use publicly available de-identified data, so our study was exempt from institutional review board approval. We followed the Strengthening the Reporting of Observational Studies in Epidemiology (STROBE) reporting guidelines for cohort studies.

## RESULTS

In Figure 1, we present estimates for percent vaccinated in each period. In March, low SES communities (23.01%; 95% CI: 20.25% - 25.76%) had significantly lower percent vaccinated than high SES communities (34.73%; 95% CI: 32.19% - 37.27%). In April, low SES communities (35.79%; 95% CI: 31.55% - 40.00%) had significantly lower percent vaccinated than high SES communities (51.65%; 95% CI: 48.44% - 54.85%). In May, low SES communities (45.65%; 95% CI: 40.49% - 50.81%) had significantly lower percent vaccinated than high SES communities (60.46%; 95% CI: 57.82% - 63.10%).

**Figure 1.**
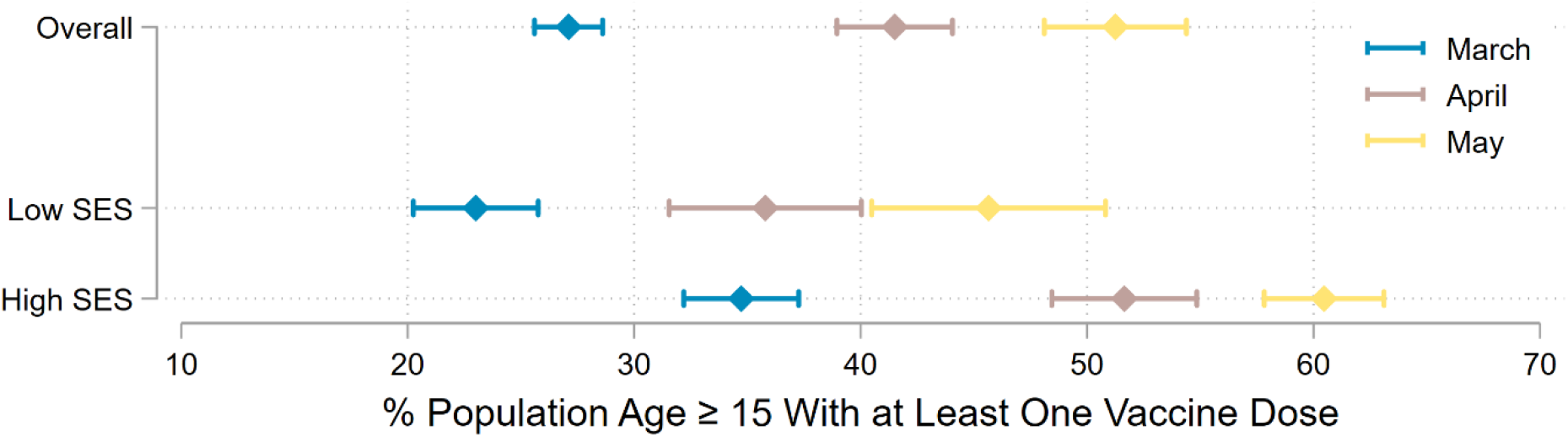
COVID-19 Vaccination by ZIP Code Population SES Composition. **Note**: This figure illustrates adjusted predictions for ZIP Codes with a given socioeconomic composition. We defined “low” and “high” as the within-city lowest quartile and highest quartile, respectively. We defined SES levels by setting all four SES variables to the same within-city quartiles within each scenario. We set other independent variables to within-city averages in each scenario.

In Figure 2, we present estimates for the change in percent vaccinated over time. Between March and April, low SES communities (12.78%; 95% CI: 11.19% - 14.38%) had significantly lower change in percent vaccinated than high SES communities (16.92%; 95% CI: 15.38% - 18.45%). Between April and May, the difference between change in percent vaccinated in low SES communities (9.86%; 95% CI: 8.77% - 10.96%) and high SES communities (8.82%; 95% CI: 7.39% - 10.25%) was not significant.

**Figure 2.**
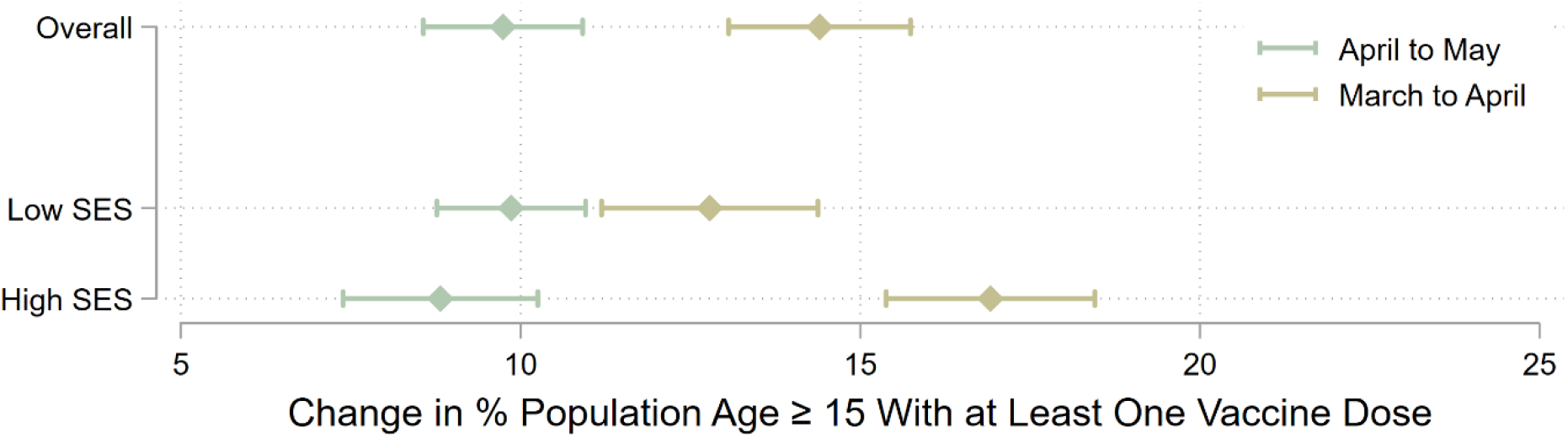
Change in COVID-19 Vaccination by ZIP Code Population SES Composition. **Note**: This figure illustrates adjusted predictions for ZIP Codes with a given socioeconomic composition. We defined “low” and “high” as the within-city lowest quartile and highest quartile, respectively. We defined SES levels by setting all four SES variables to the same within-city quartiles within each scenario. We set other independent variables to within-city averages in each scenario.

## DISCUSSION

We examined changes in COVID-19 vaccination in eight of the ten most populous cities in the United States. The percent vaccinated in low SES communities lagged high SES communities in March, April, and May. Additionally, the large gap in percent vaccinated between communities during the restricted vaccine eligibility period did not narrow when eligibility opened up in late April and early May. During the six weeks captured in our data, 64.5 million people received their first dose of a vaccine, equal to 31.2% of all vaccinated individuals as of September 1, 2021.^17^ Thus, despite the rapid and widespread reach of vaccinations during this period, large inequalities persisted.

Our work suggests that a process of cumulative disadvantage at the community – and likely individual – level is unfolding due to the COVID-19 pandemic. The same communities that suffered the highest burdens of infection and mortality from COVID before vaccines were available had lower levels of community vaccination during restricted vaccine eligibility and did not immediately close those gaps as eligibility opened up. The link between COVID-19 vaccination and community disadvantage are concerning. Persisting disparities led to geographic clusters of unvaccinated individuals, a problem that, even by itself, can spiral and may have prolonged and exacerbated the negative effects of the pandemic.^18,19^ Importantly, this continuing inequality may lead to a bifurcated recovery where advantaged communities move on from the pandemic more quickly while disadvantaged communities continue to suffer.

## Data Availability

All data produced in the present study are available upon reasonable request to the authors.

## Acknowledgments

This project was supported in part by the California Center for Population Research at the University of California, Los Angeles (UCLA) with training support (T32HD007545) and core support (P2CHD041022) from the Eunice Kennedy Shriver National Institute of Child Health and Human Development (NICHD). The content is solely the responsibility of the authors and does not necessarily represent the official views of the NICHD or the National Institutes of Health. Thanks to Meiying Li, Thalia Tom, Will Schupmann, and Yvonne Carrillo for research assistance.

## Notes

### Competing Interest Statement

The authors have declared no competing interest.

### Funding Statement

This study did not receive any funding.

### Author Declarations

This study involves only openly available human data, which can be obtained from sources listed in the paper.

## REFERENCES

1. Baden LR, El Sahly HM, Essink B, et al. Efficacy and Safety of the mRNA-1273 SARS-CoV-2 Vaccine. N Engl J Med. 2021;384(5):403–416. doi:10.1056/NEJMoa2035389

2. Thompson MG. Interim Estimates of Vaccine Effectiveness of BNT162b2 and mRNA-1273 COVID-19 Vaccines in Preventing SARS-CoV-2 Infection Among Health Care Personnel, First Responders, and Other Essential and Frontline Workers — Eight U.S. Locations, December 2020–March 2021. MMWR Morb Mortal Wkly Rep. 2021;70. doi:10.15585/mmwr.mm7013e3

3. Polack FP, Thomas SJ, Kitchin N, et al. Safety and Efficacy of the BNT162b2 mRNA Covid-19 Vaccine. N Engl J Med. 2020;383(27):2603–2615. doi:10.1056/NEJMoa2034577

4. LaFraniere S, Weiland N, Steinhauer J. Plunging Johnson & Johnson Vaccine Supply Dents State Inoculation Efforts. The New York Times. https://www.nytimes.com/2021/04/09/us/politics/johnson-johnson-coronavirus-vaccine.html. Published April 9, 2021. Accessed May 11, 2021.

5. National Academies of Sciences, Engineering, and Medicine, Division H and M, Practice B on PH and PH, Policy B on HS, Coronavirus C on EA of V for the N. Framework for Equitable Allocation of COVID-19 Vaccine. National Academies Press; 2020.

6. Wright W, Rio GMN del. A New Covid Dilemma: What to Do When Vaccine Supply Exceeds Demand? The New York Times. https://www.nytimes.com/2021/05/09/us/covid-vaccine-surplus.html. Published May 10, 2021. Accessed May 11, 2021.

7. CNN JH. All 50 states now have expanded or will expand Covid vaccine eligibility to everyone 16 and up. CNN. Published 2021. Accessed April 23, 2021. https://www.cnn.com/2021/03/30/health/states-covid-19-vaccine-eligibility-bn/index.html

8. DeBruin D, Liaschenko J, Marshall MF. Social Justice in Pandemic Preparedness. Am J Public Health. 2012;102(4):586–591. doi:10.2105/AJPH.2011.300483

9. Abedi V, Olulana O, Avula V, et al. Racial, Economic, and Health Inequality and COVID-19 Infection in the United States. J Racial Ethn Health Disparities. Published online September 1, 2020. doi:10.1007/s40615-020-00833-4

10. Clouston SAP, Natale G, Link BG. Socioeconomic inequalities in the spread of coronavirus-19 in the United States: A examination of the emergence of social inequalities. Soc Sci Med. 2021;268:113554. doi:10.1016/j.socscimed.2020.113554

11. Do DP, Frank R. Unequal burdens: assessing the determinants of elevated COVID-19 case and death rates in New York City’s racial/ethnic minority neighbourhoods. J Epidemiol Community Health. 2021;75(4):321–326. doi:10.1136/jech-2020-215280

12. McLaren J. Racial Disparity in COVID-19 Deaths: Seeking Economic Roots with Census Data. BE J Econ Anal Policy. 2021;21(3):897–919. doi:10.1515/bejeap-2020-0371

13. Reitsma MB, Claypool AL, Vargo J, et al. Racial/Ethnic Disparities In COVID-19 Exposure Risk, Testing, And Cases At The Subcounty Level In California. Health Aff (Millwood). 2021;40(6):870–878. doi:10.1377/hlthaff.2021.00098

14. Wrigley-Field E, Garcia S, Leider JP, Van Riper D. COVID-19 Mortality At The Neighborhood Level: Racial And Ethnic Inequalities Deepened In Minnesota In 2020. Health Aff (Millwood). 2021;40(10):1644–1653. doi:10.1377/hlthaff.2021.00365

15. DiRago NV, Li M, Tom T, et al. COVID-19 Vaccine Rollouts and the Reproduction of Urban Spatial Inequality: Disparities Within Large U.S. Cities in March and April 2021 by Racial/Ethnic and Socioeconomic Composition. SSRN. Published online 2021. https://papers.ssrn.com/sol3/papers.cfm?abstract_id=3918562

16. Sacarny A, Daw JR. Inequities in COVID-19 Vaccination Rates in the 9 Largest US Cities. JAMA Health Forum. 2021;2(9):e212415–e212415. doi:10.1001/jamahealthforum.2021.2415

17. CDC. COVID Data Tracker. Centers for Disease Control and Prevention. Published September 5, 2021. Accessed September 5, 2021. https://covid.cdc.gov/covid-data-tracker

18. Estep K, Greenberg P. Opting Out: Individualism and Vaccine Refusal in Pockets of Socioeconomic Homogeneity. Am Sociol Rev. 2020;85(6):957–991. doi:10.1177/0003122420960691

19. Gromis A, Liu KY. The Emergence of Spatial Clustering in Medical Vaccine Exemptions Following California Senate Bill 277, 2015–2018. Am J Public Health. 2020;110(7):1084–1091. doi:10.2105/AJPH.2020.305607

